# What role for asbestos in idiopathic pulmonary fibrosis? Findings from the IPF job exposures study

**DOI:** 10.1101/2021.03.09.21253224

**Authors:** Carl J Reynolds, Rupa Sisodia, Chris Barber, Cosetta Minelli, Sara De Matteis, Miriam Moffatt, John Cherrie, Anthony Newman Taylor, Paul Cullinan, Sophie Fletcher, Gareth Walters, Lisa Spenser, Helen Parfrey, Gauri Saini, Nazia Chaudhuri, Alex West, Huzaifa Adamali, Paul Beirne, Ian Forrest, Michael Gibbons, Justin Pepperell, Nik Hirani, Kim Harrison, Owen Dempsey, Steve O’Hickey, David Thickett, Dhruv Parekh, Suresh Babu, Andrew Wilson, George Chalmers, Melissa Wickremasinghe, Robina Coker on behalf of the IPFJES collaborators

**Author notes:** **Corresponding author** Dr Carl J Reynolds, National Heart and Lung Institute, 1b Manresa Road, London, United Kingdom, SW3, 6LR. **Authors’ Contributions:** CJR, CB, CM, MM, ANT, JC, PC made substantial contributions to the design, analysis, and interpretation of the work. CJR and RS made substantial contributions to the acquisition of data for the work. All authors contributed to revising the work critically for important intellectual content and approved the final version.

## Abstract

**Rationale:** Asbestos is posited to cause otherwise ‘idiopathic’ pulmonary fibrosis (IPF); establishing this has important diagnostic and therapeutic implications.

**Objectives:** To determine the association between occupational asbestos exposure and IPF; to investigate interaction with *MUC5B* rs35705950 genotype.

**Methods:** Multi-centre, incident case-control study. Cases (n=494) were men diagnosed with IPF at 21 United Kingdom hospitals. Controls (n=466) were age-matched men who attended a hospital clinic in the same period. Asbestos exposure was measured using a validated job exposure matrix and a source-receptor model. The primary outcome was the association between asbestos exposure and IPF, estimated using logistic regression adjusted for age, smoking and centre. Interaction with *MUC5B* rs3570950 was investigated using a genetic dominant model.

**Measurements and Main Results:** 327 (66%) cases and 293 (63%) controls ever had a high or medium asbestos exposure risk job; 8% of both cases and controls, had cumulative exposure estimates ≥ 25 fibre.ml^−1^.years. Occupational asbestos exposure was not associated with IPF, adjusted OR 1.1(95%CI 0.8-1.4; p=0.6) and there was no gene-environment interaction (p=0.2). Ever smoking was associated with IPF, OR 1.4 (95%CI 1-1.9; p=0.04). When stratifying for genotype there was significant interaction between smoking and work in an exposed job (p<0.01) for carriers of the minor allele of *MUC5B* rs3570950.

**Conclusions:** Occupational asbestos exposure alone, or through interaction with *MUC5B* rs35705950 genotype, was not associated with IPF. However, exposure to asbestos and smoking interact to increase IPF risk in carriers of the minor allele of *MUC5B* rs3570950.

Clinical trial registered with www.clinicaltrials.gov (NCT03211507).

## Introduction

Idiopathic pulmonary fibrosis (IPF) is a progressive fibrotic lung disease which in 2016 was the recorded underlying cause of death for approximately 5000 people in England and Wales.(1) The median age of onset is 70 years and the condition is more common in men (male:female ratio 1.6), manual workers, and those living in industrial areas(2), patterns that are not unique to the UK.(3)(4) The prognosis is poor, with a median survival of three years.(2) The pathophysiology of IPF is complex and believed to be the outcome of epithelial injury, with a dysregulated repair process, in a susceptible host. Several gene polymorphisms which result in a vulnerable alveolar epithelium have been characterized; they include abnormalities in mucin genes (eg *MUC5B*), surfactant protein genes, and telomerase genes (eg *TERT* and *TERC*).(3)(5)

Clinical, radiological, and histopathological findings in asbestosis and IPF are similar(6)(7). Mineralogical studies support the concept of asbestosis-IPF misclassification by revealing high fibre burdens in the lung tissue of patients diagnosed with ‘IPF’ and revision of the diagnosis to ‘asbestosis’.(8)(9)(10)(11) MUC5B is the dominant constituent of the honeycomb cysts that characterise the pattern of lung scarring, usual interstitial pneumonia (UIP), seen in both IPF and asbestosis. The strongest risk factor identified in IPF to date is the *MUC5B* promoter variant rs35705950, which increases airway expression of MUC5B(12)(13) and is also associated with increased risk of asbestosis.(14) Toxicological studies show that asbestos exposure results in production of IL-1, a key proinflammatory cytokine in IPF and a potent stimulus for MUC5B expression.(15)

In the UK, IPF mortality correlates strongly with mesothelioma mortality and lagged historic asbestos imports, ecological patterns that led Barber and his colleagues to hypothesize that “a proportion of IPF mortality is in fact due to unrecognized asbestos exposure”.(16) Occult occupational asbestos exposure as a cause for otherwise ‘idiopathic’ pulmonary fibrosis has been an open question for at least 30 years(17) and is brought to the fore in countries such as Brazil, Russia, India, and China which use large quantities of asbestos, and by the continuing global rise in asbestos-related and IPF mortality rates.(18) While occupational dust exposures in IPF have been examined by several previous case control studies, recently reviewed in a joint ATS-ERS statement(19), none has focused on assessment of asbestos exposure. The IPF job exposures study (IPFJES) addresses the question of a role for asbestos exposure in IPF by means of a job exposure matrix based on occupational proportional mortality ratios for pleural mesothelioma.(20), a validated source-receptor model(21), and examination of gene-environment interactions.

## Method

IPFJES is a multi-centre, incident case-control study. Twenty-one hospitals in England, Scotland, or Wales were selected on the basis of having a specialist IPF service, geographic dispersion and personal contacts.

Cases were men who were first diagnosed with IPF at a collaborating hospital between 01/02/2017 and 01/10/2019. The diagnosis was made at a local multi-disciplinary team meeting (MDT) using standard criteria based on clinical features, high-resolution computed-tomography (HRCT) and, when necessary, lung biopsy.(22)

At each hospital an outpatient clinic (respiratory or other) was randomly selected to serve as a source for recruitment of controls. If the clinic selected was unsuitable (it not proving possible to recruit four controls over the course of four clinics) a further random selection was made. As for cases, controls were men who attended the selected outpatient clinics between 01/02/2017 and 01/10/2019. They were frequency-matched to cases on age using five (sometimes ten) year age brackets and recruited in a 1:1 ratio to cases to achieve a predefined recruitment target.

Men unable to give informed consent or who had worked outside the UK for ≥ 1 year (not including work in the armed forces or merchant navy) were excluded. Participants were recruited by local research teams who completed a case report form and collected a blood sample which were collated centrally.

A trained interviewer (RS or CR) who was unaware of the case status of participants, administered a structured questionnaire by telephone, using a bespoke web application, to collect information on lifetime occupations, smoking, and dyspnoea (modified Medical Research Council - mMRC - dyspnoea questionnaire). For each occupation, we recorded job title, job tasks, employer and dates of employment. Occupations were automatically coded to a UK standardised occupational classification (SOC90).

Coded jobs were used to assign asbestos exposure risk through a job exposure matrix derived from occupational proportional mortality ratios for pleural mesothelioma.(20) For participants who recalled work with asbestos a detailed assessment of each work task was recorded to estimate total fibre.ml^−1^.year asbestos exposure using a validated source-receptor model.(23)(21)

For the primary analysis we used multiple logistic regression to analyse ‘any’ vs ‘no’ asbestos exposure and categories of exposure risk adjusting for age, smoking status, and recruiting centre as part of a prespecified analysis plan (clinicaltrials.gov:NCT03211507). In a secondary analysis we used multiple logistic regression to investigate gene-environment and gene-environment-environment interactions. We used a dominant rather than additive model in our interaction analysis to avoid model instability arising from the small number of participants with genotype TT.

We undertook further analyses to investigate the importance of era by analysing only jobs that ended before 1980, minimum duration in a job by analysing only jobs with a duration of ≥5 years, and cumulative ‘dose’ by multiplying duration in a job in years by a risk category weighting (office/low risk industrial 0, medium risk industrial 1, high risk construction/non-construction 2). To investigate the possibility of geographic confounding we analysed participants who lived within 10km of their recruiting hospital.

Further details are provided in the online supplement.

## Results

We recruited 516 cases and 511 controls. Twenty two cases (4%), and 45 controls (9%) were withdrawn because they no longer wished to take part in the study, they did not respond after we called them on three occasions, or we were notified that they had died before interview. The remaining 960 participants (494 cases, 466 controls) comprise the study sample.

The median year of birth and age were 1943 and 76 years for cases and 1945 and 74 years for controls (table 1). Most participants reported their ethnicity as white. 76% of cases and 70% of controls had ever smoked and only 7% of cases and 8% of controls were from higher professional socio-economic groups.

**Table 1:** Participant characteristics (N=960)

| Characteristic | Cases(N=494) | % | Controls(N=466) | % |
| --- | --- | --- | --- | --- |
| Median age in years<br>(interquartile range) | 76 (71-81) |  | 74 (69-79) |  |
| Ethnicity |  |  |  |  |
| White | 479 | 97 | 449 | 96 |
| Asian/Asian British | 11 | 2 | 8 | 2 |
| Black/African | 2 | 0 | 7 | 2 |
| Mixed/Other | 2 | 0 | 2 | 0 |
| Social class* |  |  |  |  |
| 1 | 36 | 7 | 39 | 8 |
| 2 | 57 | 12 | 61 | 13 |
| 3 | 78 | 16 | 74 | 16 |
| 4 | 50 | 10 | 51 | 11 |
| 5 | 94 | 19 | 97 | 21 |
| 6 | 114 | 23 | 89 | 19 |
| 7 | 65 | 13 | 55 | 12 |
| Smoking |  |  |  |  |
| Current smoker | 10 | 2 | 30 | 6 |
| Ever smoked | 373 | 76 | 327 | 70 |
| median pack years<br>(interquartile range) | 21 (10-38) |  | 21 (9-36) |  |
| mMRC† |  |  |  |  |
| 0 | 35 | 7 | 254 | 55 |
| 1 | 94 | 19 | 65 | 14 |
| 2 | 165 | 33 | 80 | 17 |
| 3 | 172 | 35 | 65 | 14 |
| 4 | 28 | 6 | 2 | 0 |
| rs35705950 genotype |  |  |  |  |
| GG | 152 | 38 | 327 | 77 |
| GT | 212 | 54 | 91 | 22 |
| TT | 31 | 8 | 5 | 1 |

\*Participants were assigned to National Statistics Socio-Economic analytic classes (NS-SEC) provided by the Office of National Statistics on the basis of their occupations. The seven classes used were: 1. Higher managerial, administrative and professional occupations 2. Lower managerial, administrative and professional occupations 3. Intermediate occupations 4. Small employers and own account workers 5. Lower supervisory and technical occupations 6. Semi-routine occupations 7. Routine occupations. Further details are provided in the online supplement.
†mMRC (modified Medical Research Council) dyspnoea scale. 0. Not troubled with breathlessness except with strenuous exercise 1. Troubled by shortness of breath when hurrying on the level or walking up a slight hill 2. Walks slower than people of the same age on the level because of breathlessness or has to stop for breath when walking at own pace on the level 3. Stops for breath after walking about 100 yards
or after a few minutes on the level 4. Too breathless to leave the house or breathless when dressing or undressing.(48)

All cases had a CT thorax which was reported as showing definite UIP in 266 (54%), possible UIP in 216 (44%), or ‘other’ in 12 (2%) patients. Nine cases (2%) had a lung biopsy because the CT was non-diagnostic; each of these was reported as compatible with definite UIP. Cases were more breathless than controls as measured by mMRC dyspnoea scale.

Cases and controls had a mean (sd) of 4.6 (2.4) and 4.2 (2.2) jobs respectively. Median (IQR) duration of a job for cases and controls was 5 (2-13) and 5 (2-14) years respectively. Three hundred and twenty six (66%) cases and 292 (63%) controls had ever had a high or medium asbestos exposure risk job (table 2) and did so for a median (IQR) of 25 (8-42) and 20 (6-41) years respectively. The number of years worked in a medium or high risk job did not differ significantly between cases and controls. (see Figure 1).

**Table 2:** Occupational asbestos exposure in cases and controls (N=960)

| Exposure* | Cases(N=494) | % | Controls(N=466) | % |
| --- | --- | --- | --- | --- |
| ever asbestos exposed | 327 | 66 | 293 | 63 |
| high-risk non-construction | 65 | 13 | 51 | 11 |
| high-risk construction | 138 | 28 | 126 | 27 |
| medium risk industrial | 124 | 25 | 116 | 25 |
| low risk industrial | 94 | 19 | 97 | 21 |
| office | 73 | 15 | 76 | 16 |
|  | N=122 | 25 | N=107 | 23 |
| median fibre.ml<sup>-1</sup>.years (interquartile range) | 6 (0-63) |  | 5 (0-60) |  |
| ≥100 fibre.ml<sup>-1</sup>.years | 27 | 5 | 24 | 5 |
| ≥50 fibre.ml<sup>-1</sup>.years | 34 | 7 | 29 | 6 |
| ≥25 fibre.ml<sup>-1</sup>.years | 40 | 8 | 35 | 8 |

\*Categories of occupational asbestos exposure risk were defined on the basis of occupational proportional mortality ratios for mesothelioma(20) and ever asbestos exposed was defined as ever having had a high or medium asbestos exposure risk job. Fibre.ml<sup>-1</sup>.year asbestos exposure estimates were calculated for participants who recalled working with asbestos using a source-receptor model.(23)(21)

**Figure 1.**
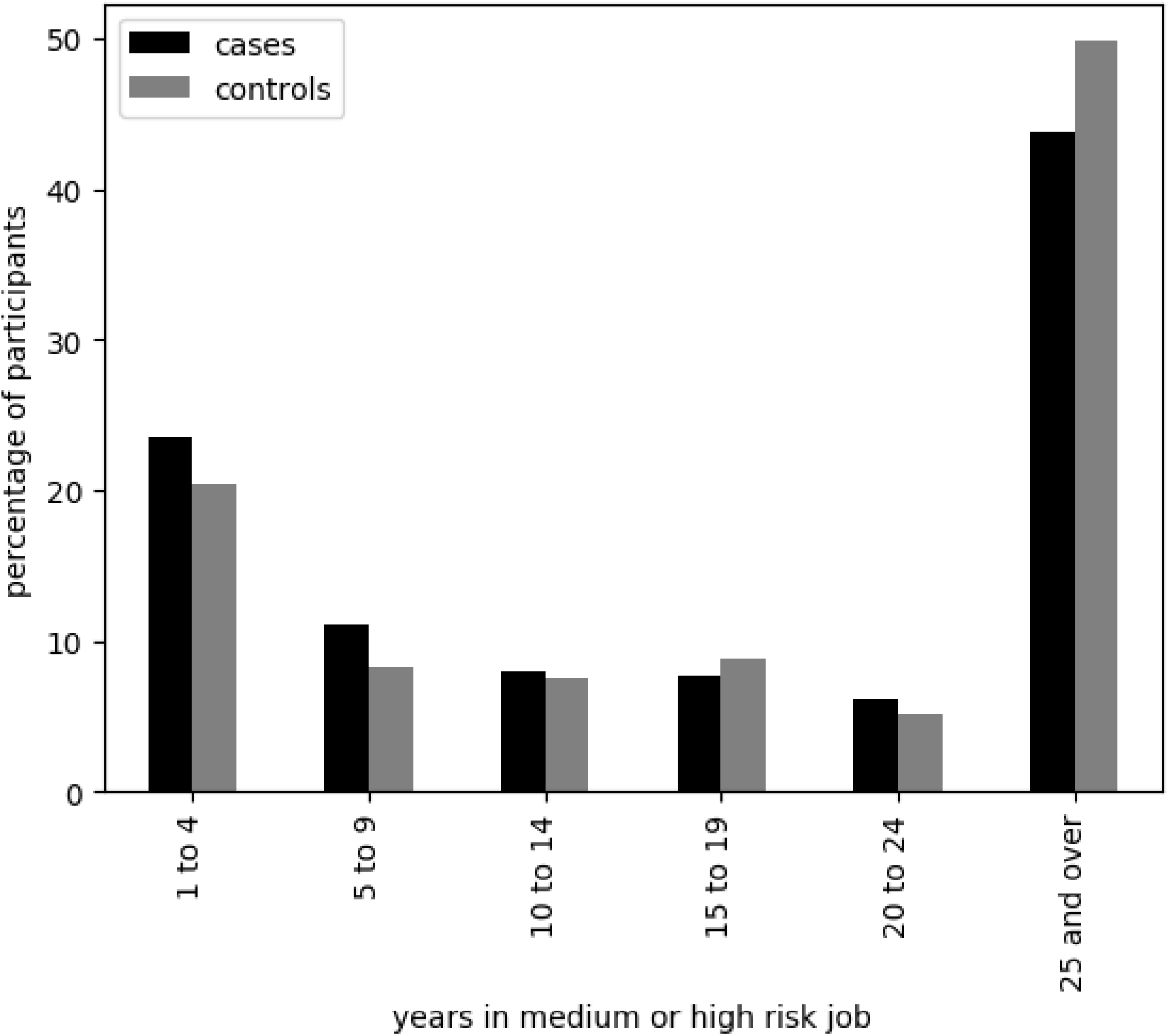
Years spent in medium or high risk jobs for all participants (N=960). A high or medium asbestos exposure risk job was defined on the basis of occupational proportional mortality ratios for mesothelioma.(20)

A total of 457 asbestos exposed job tasks were recalled in sufficient detail to permit a fibre.ml^−1^.years estimate of exposure for 229 individual participants (122, 25% of cases and 107, 22% controls). Forty (33%) cases and 35 (32%) controls, equating to approximately 8% in each group, had cumulative estimates exceeding 25 fibre.ml^−1^.years.

Fibre.ml^−1^.years exposure assessments showed reasonable agreement with those made by an independent assessor (JC) for a validation sample of low, medium, and high exposure assessments (see Figure E2 in the online data supplement).

Table 3 shows the results of an adjusted multiple regression analysis which confirmed that ever having had a high or medium asbestos exposure risk job was not associated with IPF, OR 1.1(95%CI 0.8-1.4; p=0.6). A history of ever smoking was significantly associated with an increased risk of disease, OR 1.4 (95%CI 1-1.9, p=0.04).

**Table 3:** Occupational asbestos exposure, smoking, and IPF (N=960)

| Exposure* | Adjusted OR<sup>2</sup> (95%CI;p-value) |
| --- | --- |
| ever asbestos exposed | 1.1(0.8-1.4;0.60) |
| high-risk non-construction | 0.9(0.6-1.6;0.8) |
| high-risk construction | 1.1(0.7-1.7;0.73) |
| medium risk industrial | 0.9(0.6-1.5;0.78) |
| low risk industrial | 0.9(0.6-1.4;0.57) |
| office | 1 |
| ever smoked | 1.4(1-1.9;0.04) |
| interaction model (asbestos*smoking) |  |
| ever asbestos exposed | 0.7(0.4-1.2;0.20) |
| ever smoked | 1.0(0.6-1.6;0.83) |
| ever asbestos exposed and ever smoked interaction | 1.9(1-3.6;0.04) |

\*Categories of occupational asbestos exposure risk were defined on the basis of occupational proportional mortality ratios for mesothelioma(20) and ever asbestos exposed was defined as ever having had a high or medium asbestos exposure risk job.
†Adjusted for age, centre, and smoking; smoking was not adjusted for when it was the exposure under consideration.

Eight hundred and eighteen (85%) of the 960 participants were genotyped for MUC5B rs3570950; 90 samples remain to be genotyped, because of laboratory closure due to the COVID-19 pandemic, while 52 participants did not provide a sample. Being heterozygous (GT) had an adjusted OR of 4.8 (95%CI 3.5-6.8; p<0.01) for disease while being homozygous (TT) had an adjusted OR of 12.0 (95%CI 4.5-32.5, p<0.001). There was no evidence of interaction between MUC5B rs3570950 genotype and ever having a high or medium asbestos exposure risk job, OR 1.5 (95%CI 0.8-2.7; p=0.2). When stratifying for genotype there was a statistically significant interaction between smoking and having ever worked in a high or medium asbestos exposure risk job, adjusted OR for interaction 5.0 (95%CI 1.7-15; p<0.01) for carriers of the minor allele of *MUC5B* rs3570950 (GT or TT) but not for the wild-type GG, OR 1.0 (0.4-2.4, p=0.92).(see Table 4)

**Table 4:** Occupational asbestos exposure, smoking, genotype, and IPF; interaction terms and stratified analysis (genotyped participants only, N=818)

| Exposure* | Adjusted OR† (95%CI;p-value) |
| --- | --- |
| interaction model (asbestos*smoking) |  |
| ever asbestos exposed | 0.7(0.4-1.2;0.25) |
| ever smoked | 1.0(0.6-1.6;0.95) |
| ever asbestos exposed and ever smoked interaction | 1.9(1-3.5;0.06) |
| interaction model (asbestos*genotype) |  |
| ever asbestos exposed | 1.0(0.7-1.5;0.94) |
| genotype | 3.6(2.3-5.8;0.01) |
| ever asbestos exposed and genotype | 1.5(0.8-2.7;0.2) |
| stratified interaction model (asbestos*smoking) |  |
| GG only |  |
| ever asbestos exposed | 1 (0.5-2.2;0.96) |
| ever smoked | 1.3(0.7-2.6;0.46) |
| ever asbestos exposed and ever smoked interaction | 1(0.4-2.4;0.93) |
| GT or TT |  |
| ever asbestos exposed | 0.5(0.2-1.3;0.13) |
| ever smoked | 0.6(0.3-1.5;0.28) |
| ever asbestos exposed and ever smoked interaction | 5(1.7-15;0.01) |

\*Ever asbestos exposed was defined as ever having had a high or medium asbestos exposure risk job, defined on the basis of occupational proportional mortality ratios for mesothelioma.(20) Genotype of *MUC5B* rs35705950, T is the minor allele.
†Adjusted for age +/- smoking, smoking was not adjusted for when it was
considered as an exposure. Centre was not adjusted for in this analysis because numbers were too small for one centre. Analysis limited to the 20 centres which did have sufficient numbers showed that adjusting for centre did not significantly change our results.

Further analyses suggested that era, duration of job, cumulative ‘dose’, and distance from recruiting centre, did not alter the observed associations between asbestos exposure and IPF risk. Further details are provided in the online supplement.

## Discussion

We undertook a case-control study to investigate historic occupational asbestos exposure as a potential risk factor for IPF in men and identified a significant three-way gene-asbestos-smoking interaction. For individuals carrying the *MUC5B* rs3570950 minor allele, IPF cases were five times more likely than control subjects to report a combined history of cigarette smoking and work for at least one year in a high/medium risk asbestos exposure job – a relationship that was not seen with either environmental risk factor alone.

Pulmonary fibrosis is an age-related disease caused, it is assumed, by epithelial injury in individuals with appropriate genetic susceptibility, which may or may not have an identifiable cause.(24) Polymorphism of the *MUC5B* promoter allele is a strong genetic risk factor for a range of fibrotic lung diseases including IPF(25), asbestosis(14), chronic hypersensitivity pneumonitis(26), and rheumatoid arthritis associated ILD.(27) In keeping with previous findings, the minor allele frequency in our study was significantly higher in IPF cases (35%) than controls (12%) and was strongly associated with disease in an allele dose-dependent fashion. A history of cigarette smoking, another established risk factor in IPF(28), was also significantly more common among cases than controls (76% versus 70%), with an adjusted odds ratio of 1.4. The prevalence of ever smoking was very similar to that reported from other UK studies of IPF.(29)

Given the strong ecological association between IPF mortality and historic UK asbestos imports(16), we carried out detailed interviews to examine a potential causative link between MDT-diagnosed IPF and previous occupational asbestos exposure. To avoid sole reliance on patient recall, which is unreliable(30), we used a simple job exposure matrix based on occupational proportional mortality ratios for mesothelioma, and (where possible) a validated source-receptor model. We found that working for at least a year in a high/medium risk asbestos exposure job was common in both cases (66%) and controls (63%) with no significant difference between the two in terms of social class, work duration or cumulative lifetime exposures. These findings were unaffected by adjustment for age, recruiting centre and smoking. Similarly, where an estimate was possible, there were no differences in quantified exposures. These findings indicate that, at least in UK men, there is no overall association between occupational asbestos exposure and risk of IPF.

The question of whether some cases should more properly be labelled as asbestosis naturally arises but we note that 8% of both cases and controls had estimated cumulative asbestos exposures in excess of 25 fibre.ml^−1^.years, the Helsinki criteria exposure threshold at which cases of asbestosis may occur.(31) Thus, in this generation of British men with interstitial fibrosis, a history of heavy asbestos exposure is common but no more so than in other men attending hospital.

A diagnosis of ‘asbestosis’ is made in patients with UIP who have had a substantial exposure to asbestos However, what constitutes a sufficient exposure to cause asbestosis and, in particular, at what level of cumulative exposure the risk of pulmonary fibrosis is more than double the risk in the general population, have not been formally investigated. In this study the level of asbestos exposure was not different in the the cases of UIP from the control group. No case of UIP could therefore be confidently attributed to asbestos exposure and therefore in no case could a diagnosis of asbestosis be logically made.

Prior to our study, seven case-control studies had not found self-reported occupational asbestos exposure to be a significant risk factor in IPF(17)(32)(33)(34)(35)(36)(37). In contrast, Abramson et al(38) recently published a large IPF case-control study from Australia that did find a significant association, both for self-reported asbestos exposure and cumulative estimates quantified by an asbestos job exposure matrix. Interestingly, although that study reported very low cumulative asbestos exposures in cases (mean 0.23 fibre.ml^−1^.years), a dose-response relationship between exposure and disease was still apparent. Our study found that the median duration of work in a high/medium risk asbestos exposure job for MDT-diagnosed IPF cases was 20 years, with 57% of patients having worked for at least five years in a medium risk job or at least one year in a high risk job. One in four IPF cases were able to recall previous occupational asbestos exposure, and of these, a third had had an estimated lifetime exposure in excess of 25 fibre.ml^−1^.years.

Genetic susceptibility to IPF is complex, and a limitation of our study was that we were unable to examine gene-environment interactions for the other polymorphisms associated with increased risk of disease; in addition, the COVID pandemic meant that not all the cases could be genotyped. We assessed occupational asbestos exposure in 494 male cases, more than in the largest previous study.(38) We chose hospital-based, rather than population, controls, as a more valid basis for comparison with cases of IPF diagnosed in hospital, the controls thus being representative of the source population of the cases.(39) The choice had the added advantage of a higher response rate than would be anticipated for population-based controls; just 28% of community controls responded in a recent UK IPF case-control study(40) while hospital controls generally have higher response rates.(41) Response rates are associated with socioeconomic status and there is a tendency for more deprived socioeconomic groups to be under-represented and more affluent groups over-represented in population control samples.(20) This risks selection bias, particularly in occupational studies, and the risk is greater with lower response rates.(42)

The study’s design and pre-specified analyses were pre-registered. During the asbestos exposure assessment process, the assessors were unaware of case-status and two validated means of assessing UK asbestos exposure were used, in both groups identically, to permit quantitative and semi-quantitative analysis. Our exposure estimates based on job title are very close to those from a recent, UK mesothelioma case-control study using the same method, in which 65% of male general population controls aged 37-79 had ever worked in an occupation at high or medium risk for asbestos exposure.(20) The estimates derived from the source-receptor model were dependent on participants recalling work with asbestos. While this may be a relatively insensitive measure we do not think it will have introduced any important bias; indeed, since cases were probably more likely to have been questioned about asbestos exposure prior to our study, our estimates may have been biased away from the null. A limitation is that we lack comprehensive data on participation rates. While collaborating hospitals were provided with screening logs and asked to report monthly the number of eligible participants identified, approached, and recruited, most found this difficult to organize. Figures from the three centres that did provide detailed participation rates suggest an overall participation rate of approximately 90% for cases and 85% for controls.

Our study is the first to investigate the interaction between established environmental and genetic risk factors for pulmonary fibrosis, a combination of historic occupational asbestos exposure, previous cigarette smoking and a *MUC5B* polymorphism. After stratifying for genotype, we found a significant three-way interaction between having ever smoked and having ever worked for at least a year in a high/medium asbestos exposure risk job (OR 5.0) for carriers of the minor allele of *MUC5B* rs3570950 (GT or TT). This triple combination of risk factors was present in 35% of the IPF cases who were genotyped. One interpretation of these findings is that in genetically susceptible individuals, chronic exposure to a combination of asbestos fibres and cigarette smoke results in inflammation and epithelial injury sufficient to result in pulmonary fibrosis. This model has biological plausibility since the *MUC5B* promoter variant is associated with overexpression of *MUC5B* leading to mucociliary dysfunction and retention of inhaled particles.(13)(43) In mouse studies, both cigarette smoke and asbestos exposure increase the production of reactive oxygen species that are thought to be important in the pathogenesis of pulmonary fibrosis.(44) Asbestos exposure and smoking also activate the NLRP3 inflammasome resulting in increased IL-1 release, a potent stimulus for increased *MUC5B* expression.(15)(45)(46)(47)

Overall, we find no evidence that occupational asbestos exposure alone is associated with IPF. This highlights the inherent difficulties that ILD MDTs face in terms of correctly differentiating patients with IPF and asbestosis; most men in their 70s in the UK who attend hospital with IPF have worked for prolonged periods in high or medium risk asbestos exposure jobs. That such a level of exposure is no more common in men with fibrosis than in others attending hospital suggests that making a diagnosis of asbestosis on the basis of an exposure history alone is, at best, an implicit acknowledgment of host susceptibility.

## Data Availability

aggregate data may be available on request

http://ipfjes.org

## Acknowledgments

The authors gratefully acknowledge Imperial College London, the National Institute for Health Research, the Wellcome Trust, the individuals who participated in the study that contributed to this work, and the clinical and research staff at each site for their efforts.

