## Supplementary material for "What role for asbestos in idiopathic pulmonary fibrosis? Findings from the IPF job exposures study": Online supplement

### Ethical approval

The study was approved by the NHS Health Research Authority (IRAS project ID 203355, REC reference 17/EM/0021) and the sponsor was Imperial College London.

#### Power calculation

Prior data indicated that the probability of occupational asbestos exposure in UK men aged 37-79 is 0.65.(20) If the true OR for disease in exposed men is 1.5, we calculated we would need to recruit 460 cases and controls, with power of 80% and significance threshold 0.05; our planned sample size included a margin for model stability and incomplete data. In a planned secondary analysis we investigated gene-environment interaction. The global minor allele frequency of MUC5B rs35705950 is 0.05; with an estimated prevalence of IPF of 20/100000 and with ORs of 1.5 for asbestos exposure and of 6.8 for rs35705950, 460 cases would be required to detect a minimum interaction OR of 5.0.

#### Genotyping

DNA was extracted from whole blood samples using a Nucleon™ BACC3 Genomic DNA Extraction Kit (GE Healthcare). Genotypes of the *MUC5B* SNP rs35705950 were determined using TaqMan assays (Life Technologies, Carlsbad, CA) in 96-well plates, and fluorescence read using a Viia7 Sequence Detection System (Applied Biosystems).

#### Analysis code

We undertook statistical analyses using Python, SciPy, Statsmodels, and Stata (StataCorp. 2015. Stata Statistical Software: Release 14. College Station, TX: StataCorp LP).

Analysis code is available online:

<https://github.com/drcjar/ipfjes/blob/master/notebooks/8.%20ipfjes_paper_analysis.ipynb>

#### Coding socio-economic class

SOC90 coded jobs were also used to assign National Statistics Socio-Economic analytic classes (NS-SEC). The Office of National Statistics provides a lookup to assign each SOC90 code to one of eight classes:

1. Higher managerial, administrative and professional occupations
2. Lower managerial, administrative and professional occupations
3. Intermediate occupations
4. Small employers and own account workers
5. Lower supervisory and technical occupations
6. Semi-routine occupations
7. Routine occupations
8. Never worked and long-term unemployed

We then assigned each individual to a single code by calculating the median code for all of the jobs they had held.

#### Asbestos job exposure matrix

SOC90 coded jobs were used to assign patients to one of five main categories based on the highest risk job ever held(1):

1. High-risk non-construction
2. High-risk construction
3. Medium risk industrial
4. Low risk industrial
5. Office

Mapping of SOC90 code to occupational asbestos exposure categories was based on occupational proportional mortality ratios for mesothelioma (see Figure E1). Ever exposed was defined as ever having a medium or high risk asbestos exposure category job.

#### Figure E1

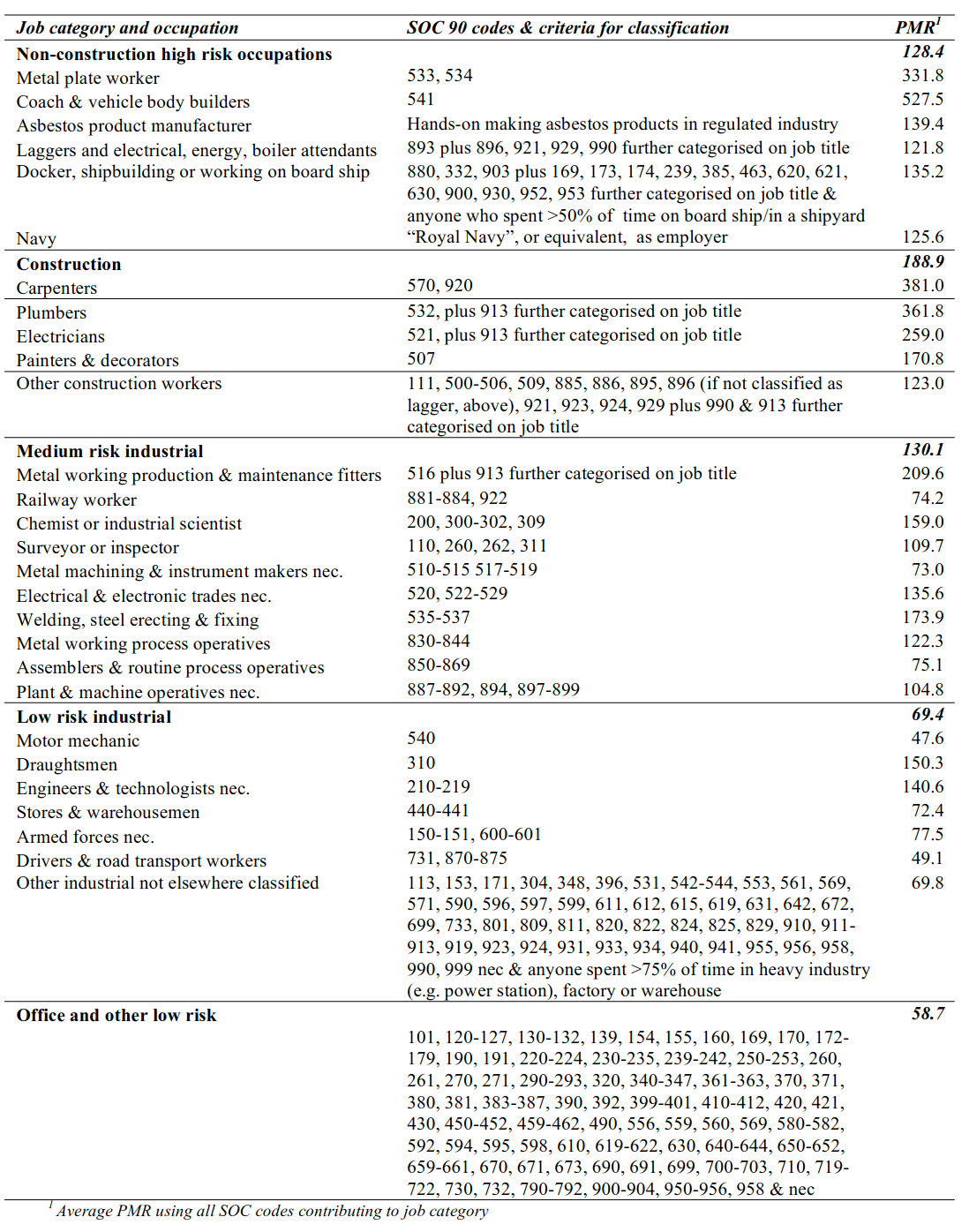
Figure E1: Classification of job categories with average national mesothelioma PMRs. Table 2.3.2 in Occupational, domestic and environmental mesothelioma risks in Britain. (1)

For analysis of categories of exposure participants were assigned to the highest risk category they ever had a job in.

#### Asbestos exposure assessment using a source receptor model

For participants who recalled carrying out work with asbestos a detailed assessment of each work task was recorded. A fibre.ml⁻¹.year asbestos exposure (AE) estimate was calculated using a source-receptor model. (2)(3)

First we calculated AE for each task as follows:

AE = E * H * LC

with parameters for the type of asbestos used (substance emission potential, E), what was done with it (activity emission potential, H), and whether there were any local exposure controls, for example wetting (local controls, LC).

AE for each task was then weighted according to the total amount of time spent performing the task and how well ventilated the room the activity was carried out in was (general ventilation parameters, D), to arrive at a task fibre.ml⁻¹.year exposure estimate.

fibre.ml⁻¹.year (job task) = AE * task_duration * (task_frequency / periodicity) * job_duration * D

Task fibre.ml⁻¹.year exposure estimates were then summed at an individual participant level to provide an overall fibre.ml⁻¹.year estimate. A random sample of five high (top 25% of values), five medium (25-75% centile), and five low (bottom 25% of values) estimates (N=15) were independently assessed by a hygiene assessment expert who was blind to participant case status. The independent assessments tended to be lower than study assessments but there was overall acceptable agreement between assessments assessed using the Bland-Altman method. (see Figure E2)

#### Figure E2

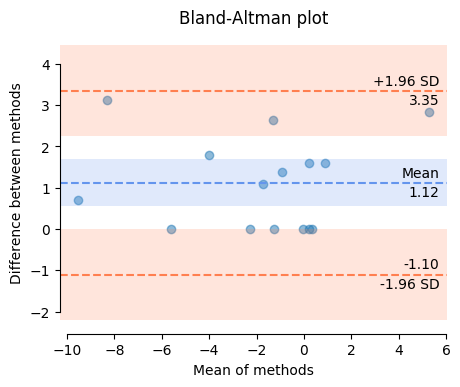

Figure E2: Independent validation of fibre.ml⁻¹.year exposure assessments. A sample of study assessments were repeated independently by an independent assessor. Difference between methods is study assessment minus the independent assessor assessment. Mean of methods is the average of the two assessments.

#### Sensitivity analyses

As a result of increasing awareness, and regulation, occupational asbestos exposure was significantly less widespread after 1980.(4) To investigate whether occupational asbestos exposure might be associated with IPF during this period we performed a sensitivity analysis by only including participants’ jobs that ended before 1980. We did not observe a significant association. We also performed sensitivity analyses limiting to jobs that started before 1980, participants born prior to 1965, and considering only jobs before age 45(5); there was no significant association between asbestos exposure and IPF for these.

#### Sensitivity analysis (limited to jobs that ended before 1980, N=771)

| Exposure* | Adjusted OR† (95%CI;p-value) |
| --- | --- |
| ever asbestos exposed | 0.9(0.7-1.3;0.66) |
| high-risk non-construction | 1.1(0.6-1.9;0.79) |
| high-risk construction | 1(0.6-1.5;0.84) |
| medium risk industrial | 0.8(0.5-1.3;0.43) |
| low risk industrial | 1(0.6-1.6;0.89) |
| office | 1 |
| ever smoked | 1.3(0.9-1.9;0.13) |
| interaction model (asbestos*smoking) |  |
| ever asbestos exposed | 0.6(0.3-1.2;0.14) |
| ever smoked | 1(0.6-1.7;0.98) |
| ever asbestos exposed and ever smoked interaction | 1.7(0.8-3.5;0.15) |

*Categories of occupational asbestos exposure risk were defined on the basis of occupational proportional mortality ratios for mesothelioma(1) and ever asbestos exposed was defined as ever having had a high or medium asbestos exposure risk job.

†Adjusted for age, centre, and smoking; smoking was not adjusted for when it was the exposure under consideration.

#### Sensitivity analysis (limited to jobs that participants spent 5 or more years in, N=957)

| Exposure* | Adjusted OR† (95%CI;p-value) |
| --- | --- |
| ever asbestos exposed | 0.9(0.7-1.2;0.59) |
| high-risk non-construction | 0.7(0.4-1.2;0.2) |
| high-risk construction | 0.9(0.6-1.4;0.77) |
| medium risk industrial | 0.7(0.5-1.1;0.89) |
| low risk industrial | 0.7(0.5-1.1;0.1) |
| office | 1 |
| ever smoked | 1.4(1-1.9;0.03) |
| interaction model (asbestos*smoking) |  |
| ever asbestos exposed | 0.7(0.4-1.1;0.11) |
| ever smoked | 1.1(0.7-1.7;0.58) |
| ever asbestos exposed and ever smoked interaction | 1.6(0.9-3;0.12) |

*Categories of occupational asbestos exposure risk were defined on the basis of occupational proportional mortality ratios for mesothelioma(1) and ever asbestos exposed was defined as ever having had a high or medium asbestos exposure risk job.

†Adjusted for age, centre, and smoking; smoking was not adjusted for when it was the exposure under consideration.

#### Sensitivity analysis (limited to participants within 10km of the recruiting hospital, N=426)

To investigate the importance of distance from recruiting hospital we estimated participants distance from the hospital by measuring the distance between their registered primary care provider (assumed to approximate residential address) and their recruiting hospital in kilometres(km) using postcode centroid data and Vincenty’s formulae, we then analysed participants for whom this distance was less than 10km.

| Exposure* | Adjusted OR† (95%CI;p-value) |
| --- | --- |
| ever asbestos exposed | 1.3(0.8-2;0.37) |
| high-risk non-construction | 1(0.4-2.3;0.96) |
| high-risk construction | 1.1(0.5-2.2;0.86) |
| medium risk industrial | 0.9(0.4-1.9;0.71) |
| low risk industrial | 0.7(0.3-1.5;0.29) |
| office | 1 |
| ever smoked | 1.3(0.8-2.1;0.35) |
| interaction model (asbestos*smoking) |  |
| ever asbestos exposed | 1 (0.4-2.4;0.95) |
| ever smoked | 1(0.4-2.3;0.99) |
| ever asbestos exposed and ever smoked interaction | 1.4(0.5-4.1;0.5) |

*Categories of occupational asbestos exposure risk were defined on the basis of occupational proportional mortality ratios for mesothelioma(1) and ever asbestos exposed was defined as ever having had a high or medium asbestos exposure risk job.

†Adjusted for age, centre, and smoking; smoking was not adjusted for when it was the exposure under consideration. One centre was excluded from the analysis because it recruited no control participants within 10km.

#### Cumulative ‘dose’ based on occupational asbestos exposure (inferred by job title)

To investigate cumulative ‘dose’ of exposure based on job title a score was assigned based on asbestos exposure risk category of each job as follows:

- high-risk non-construction : 2
- high-risk construction : 2
- medium risk industrial : 1
- low risk industrial : 0
- office : 0

Scores were then multiplied for each job by the duration in years of the job and then summed at participant level.

|  | N | mean | std | min | 25% | 50% | 75% | max |
| --- | --- | --- | --- | --- | --- | --- | --- | --- |
| cases | 494 | 24 | 30.6 | 0 | 0 | 8.5 | 39 | 126 |
| controls | 466 | 24 | 30.4 | 0 | 0 | 6.5 | 42 | 118 |

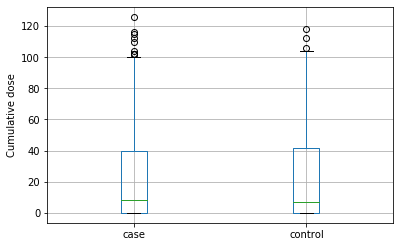

Boxplot of cumulative asbestos exposure estimates (inferred from job title) for cases and controls (N=960)

#### *MUC5B* rs35705950 minor allele frequency for genotyped cases, case subsets, and controls (N)*

|  | IPF (395) | IPF smoker (299) | IPF asbestos exposed (264) | IPF ≥25 fml.yrs (34) | IPF asbestos exposed AND smoker (213) | IPF ≥25 fml.yrs AND smoker (26) | Hospital controls (423) |
| --- | --- | --- | --- | --- | --- | --- | --- |
| GG | 152 | 112 | 100 | 11 | 76 | 9 | 327 |
| GT | 212 | 161 | 141 | 19 | 116 | 14 | 91 |
| TT | 31 | 26 | 23 | 4 | 21 | 3 | 5 |
| MAF | 35 | 36 | 35 | 39 | 37 | 38 | 12 |

*Genotype of *MUC5B* rs35705950, T is the minor allele. MAF is minor allele frequency (%). fml-yrs is cumulative fibre.ml⁻¹.years of asbestos exposure. Smoker is defined as ever smoked. Ever asbestos exposed is defined as ever having had a high or medium asbestos exposure risk job, defined on the basis of occupational proportional mortality ratios for mesothelioma.(1)
